# NeuroRehabilitation OnLine (NROL): Description of a multidisciplinary group telerehabilitation innovation for stroke and neurological conditions using the TIDieR checklist

**DOI:** 10.1101/2023.02.16.23286038

**Authors:** Suzanne Ackerley, Neil Wilson, Paul Boland, Rosemary Peel, Louise Connell

**Author notes:** Corresponding author Louise Connell, School of Sport and Health Sciences, University of Central Lancashire, UK.

## Abstract

**Background:** Providing recommended amounts of rehabilitation for stroke and neurological patients is challenging. Telerehabilitation is viable for delivering rehabilitation and an acceptable adjunct to in-person therapy. NeuroRehabilitation OnLine (NROL) was developed as a pilot and subsequently operationalised as a regional innovation embedded across four National Health Service (NHS) Trusts.

**Objective:** To describe the NROL innovation to assist future implementation and replication efforts.

**Methods:** The Template for Intervention Description and Replication (TIDieR) checklist, with guidance from the TIDieR-Telehealth extension, was used to describe NROL. The description was developed collaboratively by clinical-academics, therapists, managers, and researchers. Updated Consolidated Framework for Implementation Research domains were used to describe the context in which the innovation was delivered.

**Results:** NROL delivers online group-based real-time neurorehabilitation with technology assistance. It incorporates multidisciplinary targeted therapy and peer support to complement existing therapy. Procedures, materials and structure are detailed to demonstrate how NROL is embedded within a healthcare system. NROL uses existing NHS therapy workforce alongside dedicated NROL roles, including an essential technology support role. Selection of NROL groups is dependent on patient need. The NROL innovation is tailored over time in response to feedback. NROL described here is situated within a regional stroke and neurorehabilitation network, aligns with local and national strategies, and capitalises on an existing clinical-academic partnership.

**Conclusion:** This comprehensive description of a regional NROL innovation, and clarification of core components, should facilitate other healthcare settings to adapt and implement NROL for their context. Continuous evaluation alongside implementation will ensure maximal impact for neurorehabilitation.

## SUMMARY BOX

- The NROL innovation has been successfully integrated at a regional level to complement in-person therapy and has been comprehensively described using the TIDieR checklist.
- The core components of NROL have been clarified (Figure 1), with materials, procedures and structure detailed providing a platform to assist future implementation and replication efforts.
- Adaptations of NROL are likely to be necessary when implementing in different healthcare contexts. New iterations of NROL together with descriptions of their context should be undertaken to aid comparisons.
- Continuous evaluation alongside NROL implementation is encouraged and will ensure maximal impact for healthcare delivery.
- The model of care developed for NROL delivery may have potential use in other areas of healthcare.

## BACKGROUND

Despite a wealth of evidence that greater amounts of rehabilitation can improve outcomes(1–3), stroke and neurological patients are consistently receiving suboptimal amounts of therapy(4). Increasing access and opportunity for therapy is a critical step to addressing shortfalls in therapy amount but needs to be feasible with limited workforce. Telerehabilitation, the provision of rehabilitation remotely via telecommunication devices, offers one solution to help mitigate this challenge. It can deliver conventional in-person therapies online with equivalent outcomes(5–8) and similar attendance levels and acceptability to patients(9–11). Patients and staff report advantages in terms of saving time, energy and travel(11–13). In the UK and Ireland, clinical guidelines for stroke rehabilitation recommend telerehabilitation alongside conventional in-person therapy(14).

**Figure 1.**
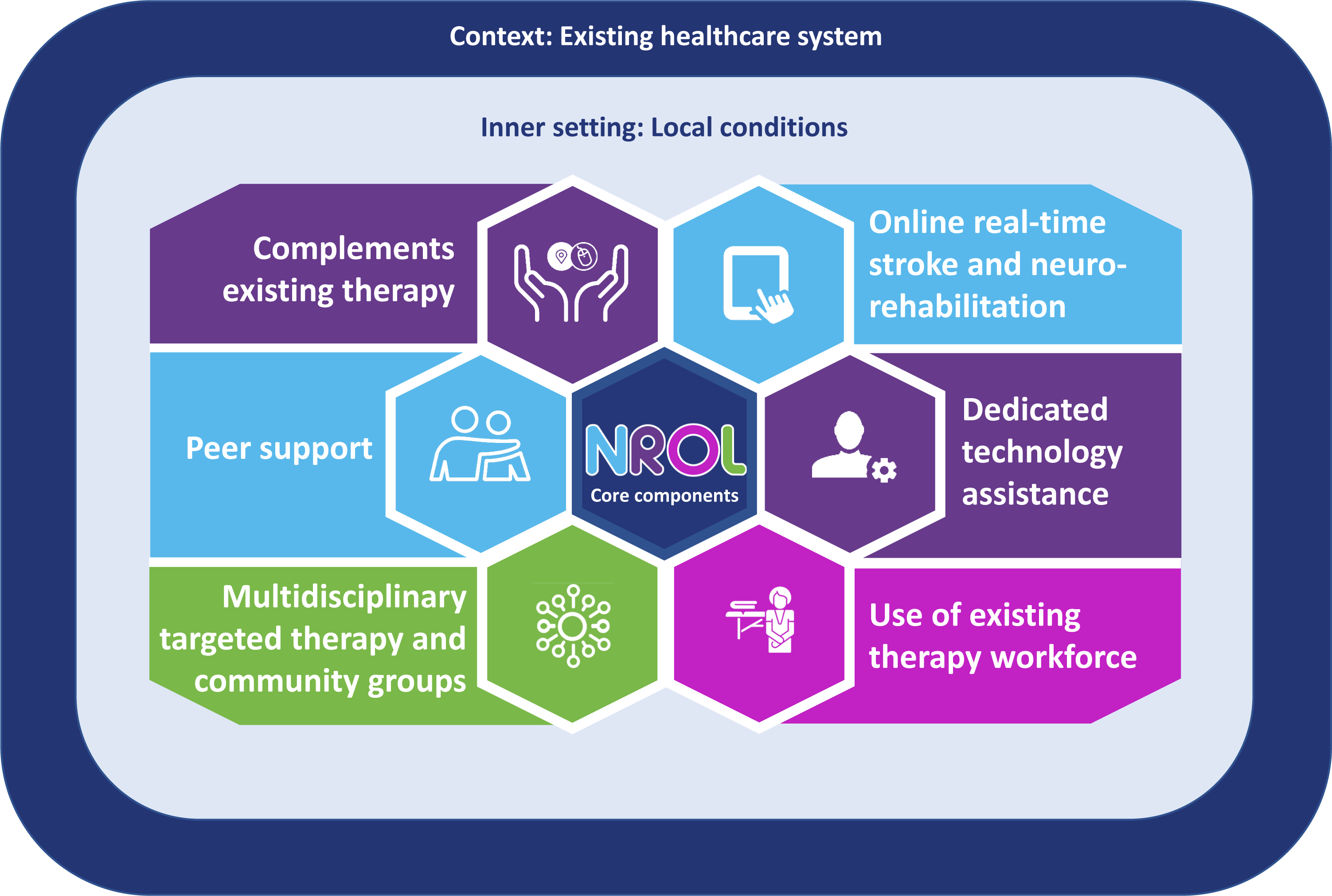
NROL core components. Six core components are identified for integration of NROL within an existing healthcare system. These components should be consistently implemented for NROL but the processes to achieve them should be adapted to fit local conditions.

A group-based real-time telerehabilitation innovation for patients with acquired brain injury was piloted in London, UK, entitled NROL (NeuroRehabilitation OnLine). This standalone version demonstrated positive impacts on patient-reported outcomes(15). NROL was subsequently adapted and operationalised within the UK National Health Service (NHS) at a single NHS Trust level yielding positive results(11). NROL was then expanded into a regional innovation involving four NHS Trusts aligning to the NHS new Integrated Care System structure to work collaboratively across regions. NROL is acknowledged as an exemplar innovation for delivering remote rehabilitation(14).

Despite the successful development of NROL, as yet it has only been described in its standalone version(15). This article describes the NROL innovation developed for regional use within an existing healthcare service, with the aim of assisting future implementation and replication efforts.

## METHODS

This article describes NROL using the 12-items of the Template for Intervention Description and Replication (TIDieR) checklist(16) and incorporates guidance from the TIDieR-Telehealth extension(17). Checklist details were developed iteratively and collaboratively, with input from staff involved in NROL implementation and evaluation including clinical-academics, therapists, managers, researchers and a patient volunteer. Initially, individual TIDieR checklists describing each NROL group in a high level of detail were produced. These informed development of NROL staff manual and standard operating procedure documents. Finally, we abstracted from these documents to develop a superordinate TIDieR checklist, aimed at describing NROL as a model of care within an existing healthcare system for application at a regional level.

Given the extensive interplay between an innovation and context, the settings in which NROL was implemented are described according to domains of the updated Consolidated Framework for Implementation Research (CFIR)(18).

### Patient and public involvement

NROL has been improved by partnership, with patients, carers and staff feedback shaping the intervention described. The NROL patient volunteer co-produced individual TIDieR checklists.

## RESULTS

### Item 1: Intervention (innovation) name

NeuroRehabilitation OnLine (NROL)

### Item 2: Why

NROL aims to enhance the rehabilitation offer for adult patients actively receiving stroke and neurorehabilitation. As part of a hybrid model of care, it utilises an online platform offering advantages to save time, energy, and travel, enabling more therapy to be delivered using existing workforce(11). Group therapy has favourable evidence(15), leveraging the benefits of peer support(19). By embedding NROL within the existing NHS system, it supports sustainable service delivery. NROL aligns with strategic priorities, such as the use of data and digital technologies in healthcare(20). Collective use of the workforce, as a provider collaborative, fosters a community of practice and shared learning(21) and also allows for a critical mass of patients to receive group therapy where impairment incidence is low.

### Item 3: What (materials) and Item 4: What (procedures)

A secure ‘NROL hub’ collaboration platform (in Microsoft (MS) Teams) was created as a repository for shared resources. Key documents include an ‘NROL standard operating procedure’ and relevant approvals (e.g., Data Protection Impact Assessment). NROL branding (i.e., logo) increases visibility and facilitates team cohesion.

The materials and procedures for NROL referral, entry, delivery and exit phases are detailed below, and outlined in an NROL process chart within the supplementary file.

#### NROL referral materials and procedures

Therapy team members identify, consent and refer suitable patients to appropriate NROL groups. They submit an ‘NROL referral form’ via the NROL hub, guided by an ’NROL staff manual’ (Supplementary file). An ’NROL patient information leaflet’ is provided to patients.

#### NROL entry materials and procedures

NROL support staff process referrals and coordinate timetabling using an NROL database (MS Excel). Prior to programme start, an NROL staff member contacts new patients to complete ’NROL outcome measures’. Measures include the EQ-5D-5L for measuring health-related quality of life(22) and the Patient Specific Functional Scale (PSFS) for measuring activity performance(23). Additionally, an NROL technology support staff member ensures that each patient has the necessary technology capability and equipment (hardware, software (MS Teams and email) and connectivity) to access NROL. An ‘NROL technology support guide’ is provided. Patients participating in physical groups receive an ’NROL physical group guide’ providing instructions on how to set up a safe exercise environment. Patients (and referrer) receive a personalised ’NROL entry email’ that outlines their 6-week programme, and group invites.

#### NROL delivery materials and procedures

Patients join NROL groups according to their personalised programme. These include targeted talking (e.g., cognition, communication, fatigue, living well) and physical (e.g., balance/mobility and upper limb) therapy groups incorporating interactive, educational, and practical elements. These groups are staffed by at least two group facilitators, often jointly by more than one discipline. Community groups are offered to all patients and include an NROL introduction and an optional weekly peer support group. Throughout the programme and during each NROL session, technology assistance is available to both patients and staff. If a patient is expected to join a group but does not attend, the NROL technology support staff member makes contact to offer assistance.

NROL sessions are delivered by staff online (MS Teams) using existing devices equipped with webcams and microphones. For physical groups, a large screen television is used for monitoring patients during exercise. The session content is developed by group facilitators based on evidence-based practice and may include discussions, demonstrations, and presentations. The NROL database is used during sessions to access and record patient information, such as attendance. A telephone is required in case of adverse events. Group facilitators ensure that clinical notes are entered for all patients. Regular NROL staff meetings and group-specific meetings are held to discuss NROL delivery.

Patients participate in NROL using their agreed device. For patients in physical groups, a large screen device is necessary. Specific equipment, such as a frame or table, may be required to ensure stability. Patients are advised to wear comfortable footwear and clothing and have access to a suitable drink. Functional task practice may necessitate additional equipment e.g., pen and paper, kitchen items. Patients should have a telephone available in case of technology issues or adverse events. If patients have pendant alarms, they are encouraged to wear them.

#### NROL exit materials and procedures

Patients (and referrer) receive an ’NROL exit email’ that provides summary information. Outcome measures are repeated, and patients are asked to complete a satisfaction survey.

#### Evaluation

Evaluation occurs concurrently with NROL delivery. Service and patient level data are sourced from the NROL database. The NROL leads undertake regular relevant analysis and summarise the data for feedback for multiple clinical and audiences.

#### Staff training

NROL evaluation findings are discussed in meetings. General NROL information is disseminated to staff across the participating trusts by NROL staff. Shared learning sessions are run to optimise knowledge translation. New staff members are introduced to NROL during their orientation. Staff can observe NROL sessions for experiential learning.

### Item 5: Who

Details of who is involved in NROL service delivery are provided in Table 1. NROL uses an existing NHS workforce alongside dedicated NROL roles for operations, administration, assistance, and technology support.

**Table 1:**
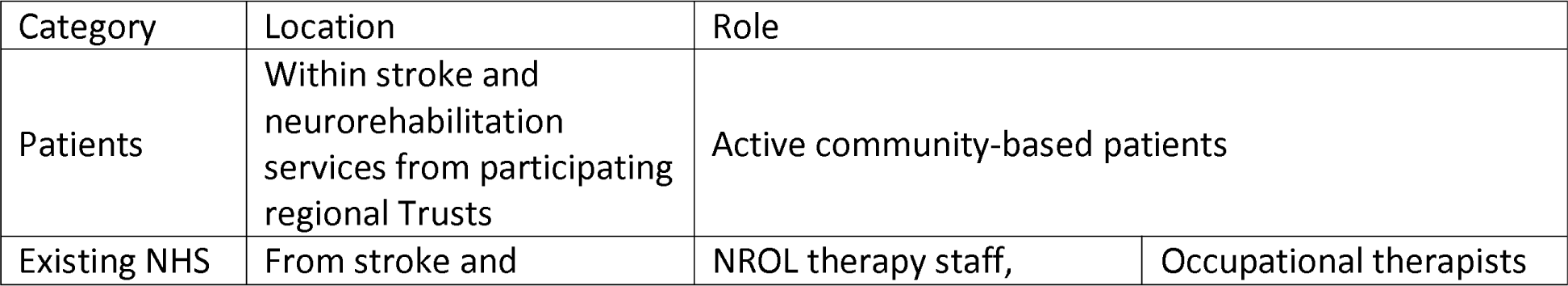
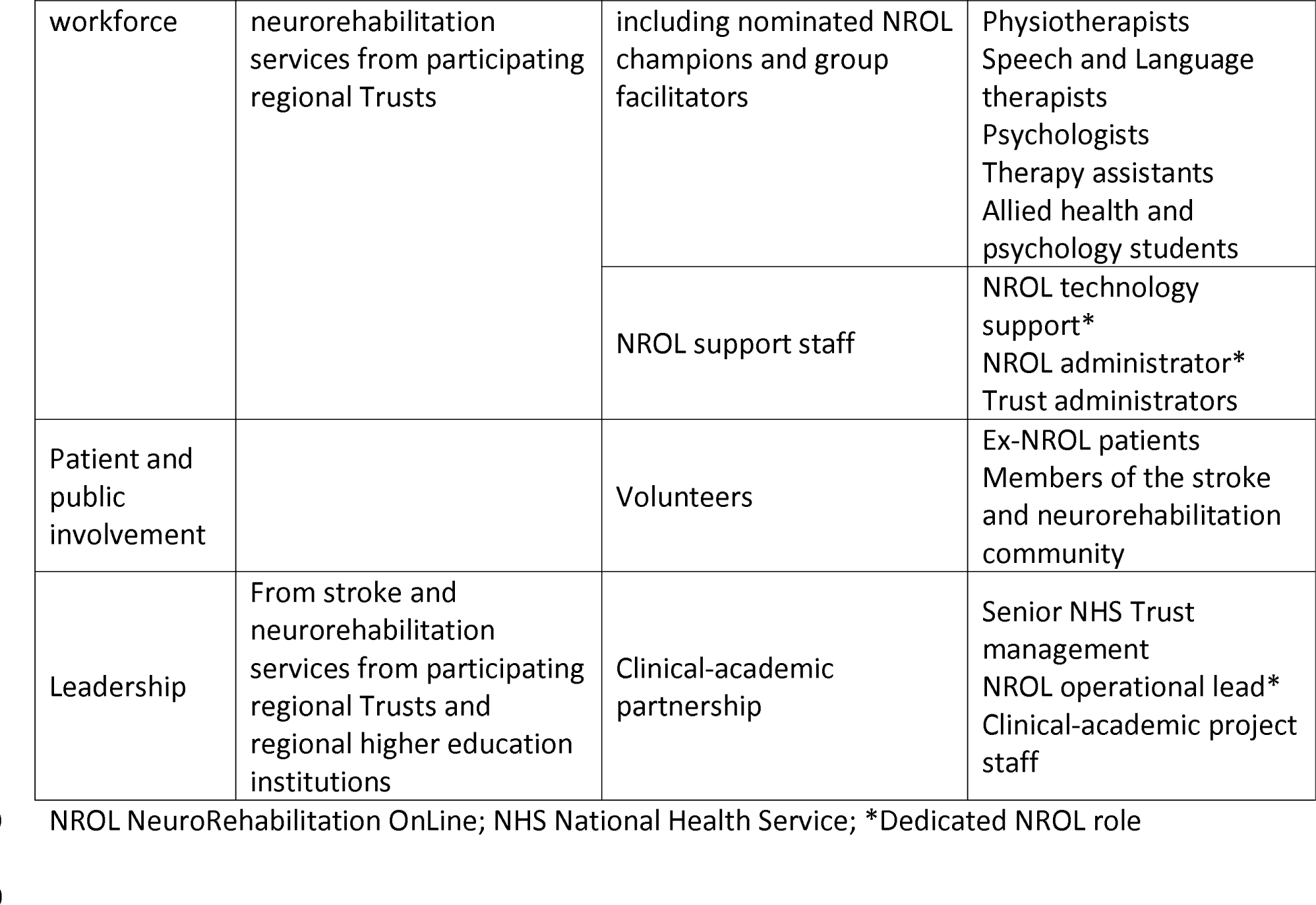
Who is involved in NROL

| Category | Location | Role |  |
| --- | --- | --- | --- |
| Patients | Within stroke and neurorehabilitation services from participating regional Trusts | Active community-based patients |  |
| Existing NHS | From stroke and | NROL therapy staff, | Occupational therapists |
| workforce | neurorehabilitation services from participating regional Trusts | including nominated NROL champions and group facilitators | Physiotherapists<br>Speech and Language therapists<br>Psychologists<br>Therapy assistants<br>Allied health and psychology students |
|  |  | NROL support staff | NROL technology support*<br>NROL administrator*<br>Trust administrators |
| Patient and public involvement |  | Volunteers | Ex-NROL patients<br>Members of the stroke and neurorehabilitation community |
| Leadership | From stroke and neurorehabilitation services from participating regional Trusts and regional higher education institutions | Clinical-academic partnership | Senior NHS Trust management<br>NROL operational lead*<br>Clinical-academic project staff |

NROL NeuroRehabilitation OnLine; NHS National Health Service; \*Dedicated NROL role

### Item 6: How (mode of delivery)

Mode of delivery overlaps with the material and procedures, see Items 3 and 4.

### Item 7: Where

Group facilitators and the NROL technology support staff member attend from private, well-lit, and quiet workspaces at different geographical locations and NHS trusts. A room with adequate space is required for demonstrating exercises for physical groups. Patients participate in NROL groups from their homes.

### Item 8: When and how much

NROL delivery is structured into recurring 6-week ‘NROL’ blocks to facilitate patient flow. An NROL introductory group runs at the start of each block. All further group sessions are scheduled for 60 minutes, with most groups run weekly. The maximum number of patients within a group is determined by the group facilitators to ensure the best experience for patients and staff (see NROL staff manual).

### Item 9: Tailoring

Tailoring is required at patient, group and block levels. Clinical reasoning should determine how NROL fits with a patient’s overall rehabilitation. Patients can attend more than one NROL block if clinically indicated, providing they remain under the active care of their stroke or neurorehabilitation team. The needs of patients in groups will be nuanced over time requiring a responsive approach. All referrals are screened by group facilitators to ensure session content is tailored. The structure of blocks, in terms of groups offered and frequency, actively considers patient need, workforce availability, and capitalises on changing staff skill sets. Resourceful use of available workforce is encouraged, such as enabling staff to deliver NROL whilst working from home or by those requiring work adjustments and involving students. NROL materials are continually edited to reflect updates.

### Item 10: Modifications

This article describes the NROL innovation modified for regional use. The core components retained from earlier iterations(11, 15) include provision of online real-time neurorehabilitation with technology assistance, incorporating multidisciplinary targeted therapy and community groups whilst embodying peer support. Adaptations for integration within an existing healthcare system have led to additional core components including delivery as an adjunct to complement existing rehabilitation and use of existing workforce. Further modifications include running NROL as recurring 6-week blocks and inclusion of patients with stroke or other neurological conditions at varying chronicity.

### Item 11: How well planned & Item 12: How well delivered (including fidelity)

Communications, resources and technology assistance are provided to optimise NROL entry and participation (see Items 3 & 4). Service data are obtained and reviewed to monitor performance.

A detailed mixed-methods evaluation of NROL within a single trust is available(11). Implementation and evaluation of the regional NROL innovation is ongoing.

### Context

The NROL innovation detailed in this article is delivered within a context that can be described using the domains of inner and outer settings(18).

The inner setting is defined as the four NHS Trusts that are situated within the Lancashire and South Cumbria Integrated Stroke and Neurorehabilitation Delivery Network that provide community-based stroke and neurorehabilitation care for the region. The region has a population of 1.8 million and covers a large geographical footprint with urban and rural settlements, and ethnic diversity. Deprivation and poor health affect many, with differences in life expectancy and quality of life varying significantly, in some neighbourhoods, healthy life expectancy is 46.5 years(24).

Aligning with national strategies and policies, the Lancashire and South Cumbria region has a vision to work collaboratively across Trusts. A challenge is that Trusts have varied service remits (stroke, neurological or both) and infrastructure (e.g., physical, staffing levels, technology systems, governance processes).

With regards to the outer setting, the impetus for starting NROL was the global pandemic, which also influenced sociocultural values of staff and patients to increase the worthiness and openness to use of remote technology. NROL also aligned with wider policy and strategies(20), benefitted from an already established clinical-academic partnership between the NHS and a university, and had funding from external sources (SameYou, NHS England).

## DISCUSSION

NROL utilises an online delivery platform, with dedicated technology assistance, to provide multidisciplinary real-time group therapy. Telerehabilitation is recognised as having a vital role to play in future healthcare delivery but as yet there are limited details of how to do this(25). To address this need, the TIDieR checklist is used to provide a comprehensive description of NROL. The actual process of completing the TIDieR was time intensive but did provide the impetus for the team to clearly describe the innovation, agreeing the core components. Consideration was given to the necessary balance of information to ensure comprehensive detail but attempting not to overwhelm. Further documentation is available within the supplementary file and by contacting the corresponding author.

Optimal adaptation of an innovation requires an understanding of the core components that cannot be changed versus the adaptable periphery that can be changed(18, 26). It is proposed that the core components identified (Figure 1) should be consistently implemented for NROL but that the processes to achieve them are adapted to fit local conditions. Examples include the use of MS Teams to deliver groups but other online platforms are available; use of recurring 6-week NROL blocks but other timings may suit other services; the number and types of targeted therapy groups will need to reflect workforce capability and capacity. It is known innovations that have adaptability are more likely to be used in clinical practice(27). Ongoing examination of the adaptive components of NROL will help discern how it can be upscaled for use in a variety of contexts.

Context is everything(28). A limitation of the TIDieR checklist is that it does not include an item on context. Arguably to understand the innovation fully and guide future adaptations an understanding of the context is required. This is because innovations are inextricably linked to the context in which they are delivered, and achieving a good fit between these is important to ensure the innovation works as intended(29). In this article, context is deliberatively reported to help situate the innovation. NROL did take resources, time and effort to implement as a regional initiative and details on the implementation will be reported elsewhere. The need for resources to enable implementation is not unexpected(13, 26) and current healthcare systems are often not set-up to facilitate this upfront effort. Influential contextual factors included leadership buy-in and commitment, a clinical-academic partnership and fit with the local and broader strategic landscape.

Inevitably NROL will continue to evolve. The TIDieR checklist does allow for reporting on modifications and tailoring. To date, the checklist has been primarily used for reporting interventions in trials(30) and there are limited examples of its use for adapted interventions over time. This article documents current NROL delivery and captures its retrospective modifications and tailoring. Going forward, transparent reporting of new iterations of NROL together with descriptions of their context should be undertaken to aid comparisons. The model of care developed for NROL delivery may have potential use in other areas of healthcare.

## CONCLUSION

A hybrid approach incorporating telerehabilitation, to complement in-person therapy, is required for a future-proof service that follows policy and guidelines. This comprehensive description of a regional NROL innovation gives an example of successful implementation within an existing healthcare system. It provides a platform for others to reduce duplication of effort and help facilitate the use of telerehabilitation in clinical practice. Adapted versions of NROL are expected when implementing in different contexts. Transparent reporting and continuous evaluation alongside NROL implementation are encouraged and will ensure maximal impact for neurorehabilitation delivery.

## Availability of data/resources

Digital copies of NROL materials are available on request by contacting Professor Louise Connell.

## Author contributions

SA and LC conceived the study. All authors (SA, LC, NW, PB, RP) were involved in the planning and conduct of the work described in the paper. SA, LC and NW drafted the initial manuscript. All authors were involved in revising the manuscript. All authors have given final approval of the version to be published and agree to be accountable for all aspects of the work.

## Data Availability

All data produced in the present study are available upon reasonable request to the authors

## Acknowledgements

We would like to express our gratitude to the individuals and services that contribute to NROL, and have contributed to this work, from East Lancashire Hospitals NHS Trust, University Hospitals of Morecambe Bay NHS Foundation Trust, Blackpool Teaching Hospitals NHS Foundation Trust and Lancashire and South Cumbria NHS Foundation Trust. Special thanks go to Ian Grimshaw for providing technological support for patients and staff, as well as supporting administrative and organisational operations. We extend our appreciation to the UCL N-ROL team for sharing their expertise and experience of online delivery and valuable discussions. Many thanks to the SameYou team for their generous fundraising and support which have made NROL possible, and to NHS England for the SQuIRe Catalyst funding to support commissioning intentions.

## REFERENCES

1. Lohse KR, Lang CE, Boyd LA. Is more better? Using metadata to explore dose-response relationships in stroke rehabilitation. Stroke. 2014;45(7):2053–8. 10.1161/strokeaha.114.004695

2. Schneider EJ, Lannin NA, Ada L, et al. Increasing the amount of usual rehabilitation improves activity after stroke: a systematic review. Journal of Physiotherapy. 2016;62(4):182–7.https://doi.org/10.1016/j.jphys.2016.08.006

3. French B, Thomas LH, Coupe J, et al. Repetitive task training for improving functional ability after stroke. Cochrane Database Syst Rev. 2016;11(11):Cd006073.DOI:10.1002/14651858.CD006073.pub3

4. Sentinel Stroke National Audit Programme. The Road to Recovery. The Ninth SSNAP Annual Report. [Internet] 2022. Available from: https://www.strokeaudit.org/Documents/National/Clinical/Apr2021Mar2022/Apr2021Mar2022-AnnualReport.aspx [accessed 30 March 2023]

5. Cramer SC, Dodakian L, Le V, et al. A Feasibility Study of Expanded Home-Based Telerehabilitation After Stroke. Front Neurol. 2020;11:611453.10.3389/fneur.2020.611453

6. Laver K, Walker M, Ward N. Telerehabilitation for Stroke is Here to Stay. But at What Cost? Neurorehabilitation and Neural Repair. 2022;36(6):331–4.10.1177/15459683221100492

7. Parrott D, Ibarra S. Randomized Controlled Trial Comparing a Telemedicine Brain Injury Coping Skills (BICS) group intervention to traditional in-person BICS for Brain Injury Patients and Caregivers. Archives of Physical Medicine and Rehabilitation. 2021;102(10):e15.https://doi.org/10.1016/j.apmr.2021.07.434

8. Khan F, Amatya B, Kesselring J, et al. Telerehabilitation for persons with multiple sclerosis. Cochrane Database Syst Rev. 2015(4):CD010508.10.1002/14651858.CD010508.pub2

9. Yang CL, Waterson S, Eng JJ. Implementation and Evaluation of the Virtual Graded Repetitive Arm Supplementary Program (GRASP) for Individuals With Stroke During the COVID-19 Pandemic and Beyond. Phys Ther. 2021;101(6).10.1093/ptj/pzab083

10. Rietdijk R, Power E, Attard M, et al. Acceptability of telehealth-delivered rehabilitation: Experiences and perspectives of people with traumatic brain injury and their carers. Journal of Telemedicine and Telecare. 2022;28(2):122–34.10.1177/1357633×20923824

11. Ackerley S, Wilson N, Boland P, et al. Implementation of neurological group-based telerehabilitation within existing healthcare during COVID-19: a mixed methods evaluation. BMC Health Services Research journal. 2023;23(1):671.

12. Buckingham S, Anil K, Demain S, et al. Telerehabilitation for people with physical disabilities and movement impairment: development and evaluation of an online toolkit for practitioners and patients. Disabil Rehabil. 2023;45(11):1885–92.10.1080/09638288.2022.2074549

13. Signal N, Martin T, Leys A, et al. Implementation of telerehabilitation in response to COVID-19: lessons learnt from neurorehabilitation clinical practice and education. New Zealand Journal of Physiotherapy. 2020;48(3):117–26.http://dx.doi.org/10.15619/NZJP/48.3.03

14. Royal College of Physicians. National Clinical Guideline for Stroke for the United Kingdom and Ireland. [Internet] 2022. Available from: https://uploads-ssl.webflow.com/62c3d8070eda8516a8ba9475/6380a7e6a9a3db745a8e8315_2023%20National%20Clinical%20Guideline%20for%20Stroke%20consultation%20document.pdf [accessed 30 March 2023]

15. Beare B, Doogan CE, Douglass-Kirk P, et al. Neuro-Rehabilitation OnLine (N-ROL): description and evaluation of a group-based telerehabilitation programme for acquired brain injury. Journal of Neurology, Neurosurgery and Psychiatry. 2021;92(12):1354.10.1136/jnnp-2021-326809

16. Hoffmann TC, Glasziou PP, Boutron I, et al. Better reporting of interventions: template for intervention description and replication (TIDieR) checklist and guide. BMJ. 2014;348(mar07 3):g1687-g.10.1136/bmj.g1687

17. Rhon DI, Fritz JM, Kerns RD, et al. TIDieR-telehealth: precision in reporting of telehealth interventions used in clinical trials - unique considerations for the Template for the Intervention Description and Replication (TIDieR) checklist. BMC Medical Research Methodology. 2022;22(1).10.1186/s12874-022-01640-7

18. Damschroder LJ, Reardon CM, Widerquist MAO, et al. The updated Consolidated Framework for Implementation Research based on user feedback. Implementation Science. 2022;17(1).10.1186/s13012-022-01245-0

19. Clark E, Maccrosain A, Ward NS, et al. The key features and role of peer support within group self-management interventions for stroke? A systematic review. Disability and Rehabilitation. 2020;42(3):307–16.10.1080/09638288.2018.1498544

20. NHS England and NHS Improvement. 2022/23 priorities and operational planning guidance. [Internet] 2022. Available from: https://www.england.nhs.uk/wp-content/uploads/2022/02/20211223-B1160-2022-23-priorities-and-operational-planning-guidance-v3.2.pdf [accessed 30 March 2023]

21. NHS England. National Stroke Service Model: Integrated Stroke Delivery Networks. [Internet] 2021. Available from: https://www.england.nhs.uk/wp-content/uploads/2021/05/stroke-service-model-may-2021.pdf [accessed 30 March 2023]

22. EuroQol Research Foundation. EQ-5D-3L User Guide. [Internet] 2018. Available from: https://euroqol.org/publications/user-guides [accessed 30 March 2023]

23. Stratford P, Gill C, Westaway M, et al. Assessing Disability and Change on Individual Patients: A Report of a Patient Specific Measure. Physiotherapy Canada. 1995;47(4):258–63.10.3138/ptc.47.4.258

24. Lancashire Independent Economic Review. Deep Dive: Health,Wealth & Wellbeing. [Internet] 2021. Available from: https://www.lancashireier.org/wp-content/uploads/2021/12/LIER_Health_Wealth_and_Wellbeing_2021_v1.pdf [accessed 30 March 2023]

25. English C, Ceravolo MG, Dorsch S, et al. Telehealth for rehabilitation and recovery after stroke: State of the evidence and future directions. International Journal of Stroke. 2022;17(5):487–93.DOI:10.1177/17474930211062480

26. Greenhalgh T, Papoutsi C. Spreading and scaling up innovation and improvement. BMJ. 2019:l2068.10.1136/bmj.l2068

27. Rogers E. Diffusion of innovations: 5th Edition. New York: Free Press; 2003.

28. Bate P. The Health Foundation: Perspectives on context. [Internet] 2014. Available from: https://www.health.org.uk/sites/default/files/PerspectivesOnContextBateContextIsEverything.pdf [accessed 30 March 2023]

29. Hawe P. Lessons from Complex Interventions to Improve Health. Annual Review of Public Health. 2015;36(1):307–23.10.1146/annurev-publhealth-031912-114421

30. Carlsson H, Rosén B, Björkman A, et al. SENSory re-learning of the UPPer limb (SENSUPP) after stroke: development and description of a novel intervention using the TIDieR checklist. Trials. 2021;22(1).10.1186/s13063-021-05375-6

