## Supplementary file for "NeuroRehabilitation OnLine (NROL): Description of a multidisciplinary group telerehabilitation innovation for stroke and neurological conditions using the TIDieR checklist"

NRO L

NeuroRehabilitation OnLine

### Lancashire and South Cumbria

### NROL Staff Manual

### Contents


**ALL ABOUT NROL**

- What is NROL?
- Overview of NROL procedures and materials
- Who is suitable for NROL?
- Consenting a patient for NROL
- Referring a patient to NROL

**NROL GROUP DESCRIPTIONS**

- GROUPS OVERVIEW
  - **Community groups:**
    - Meet and Greet
    - Café NROL
  - **Targeted therapy groups:**
    - ***Physical Groups:***
      - NROL With It
      - Upper Limb
    - ***Talking Groups:***
      - Cogs in Motion
      - Cogs in Action
      - Simply Speaking
      - Tip of the Tongue
      - Living Well
      - Fatigue Management

### ALL ABOUT NROL

#### WHAT IS NROL?

**NROL** (NeuroRehabilitation OnLine) is a **multidisciplinary group telerehabilitation innovation** that aims to increase access and opportunity to therapy for patients with stroke and other neurological conditions who are actively receiving community-based care.

NROL has 6 core components, and processes have been developed to fit them within our Lancashire and South Cumbria ISNDN. Our regional innovation involves four NHS Trusts (ELHT, UHMB, LSCFT, BTH). Patients are referred to NROL by their treating therapist.

**NROL core components:**

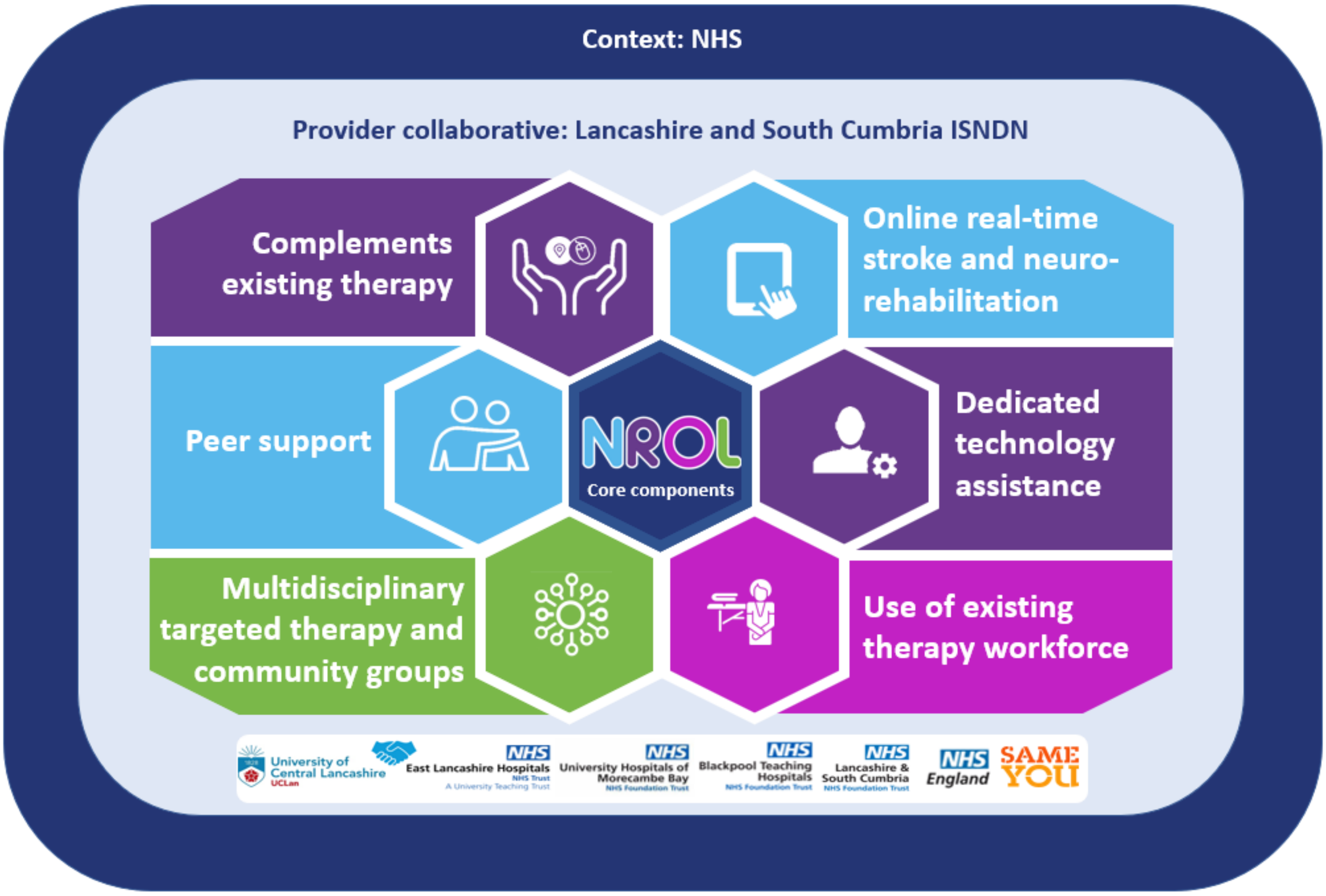


**NROL at Lancashire and South Cumbria:**

- provides **multidisciplinary targeted therapy and community groups** embodying **peer support** (see group descriptions in this manual), with relevant groups selected by the referrer
- has dedicated **technology assistance** to help patients access NROL, with in-session assistance provided
- is **delivered by therapists** from across the 4 NHS trusts
- **complements a patient's existing neurological rehabilitation**
- is delivered in recurring **6-week blocks** via the **online** platform Microsoft Teams (MS Teams)
- has patients attend from their own homes, and NROL group facilitators attend from their workspace
- the NROL team provides **online set-up, programme information and invites to each NROL patient**, with referrers b'cced into relevant NROL correspondence
- session **attendance is recorded** in patients' clinical notes
- has overall (collected by NROL team) and group-specific outcome measures (collected by therapists)

### WHO IS SUITABLE FOR NROL?

**To be included in NROL patients must:**

- ✓ be under the active care of a treating therapist in a stroke or neuro rehab team
- ✓ be willing to engage in online group therapy
- ✓ be able to communicate (if required consult with a SLT), or be supported by someone who can, in the English Language
- ✓ have a private, well-lit, quite space to join groups
- ✓ have access to an electronic device (preferably a lap-top or tablet - essential for physical groups)
- ✓ have reliable internet connectivity
- ✓ have a telephone available in case of technology issues or adverse events and should wear their pendant alarm if they have one
- ✓ meet group-specific eligibility criteria for referred groups (see group descriptions)

### OVERVIEW OF NROL PROCEDURES AND MATERIAL

NROL coordination is provided by a NROL team (NROL operational lead, therapy assistant, technology support staff member and administrator). NROL is hosted by ELHT and supported by a clinical-academic partnership with UCLan. If you have a NROL query, please contact the NROL team via.

A secure '**NROL hub**' collaboration platform provides a repository for shared NROL information and resources, including the **NROL referral form**. Please ask the NROL administrator for access via.

The key NROL procedures, and staff and patient materials, are outlined in the **'NROL process chart'** (see overleaf**)**. They depicted over four stages; Referral, Entry, Delivery and Exit, with the staff involved indicated. This chart can be used by the wider stroke and neurorehabilitation teams to help plan and organise participation for suitable patients and understand the resources available and needed from both a staff and patient perspective to support this. MS, Microsoft; IT, information technology.

**NROL process chart:**

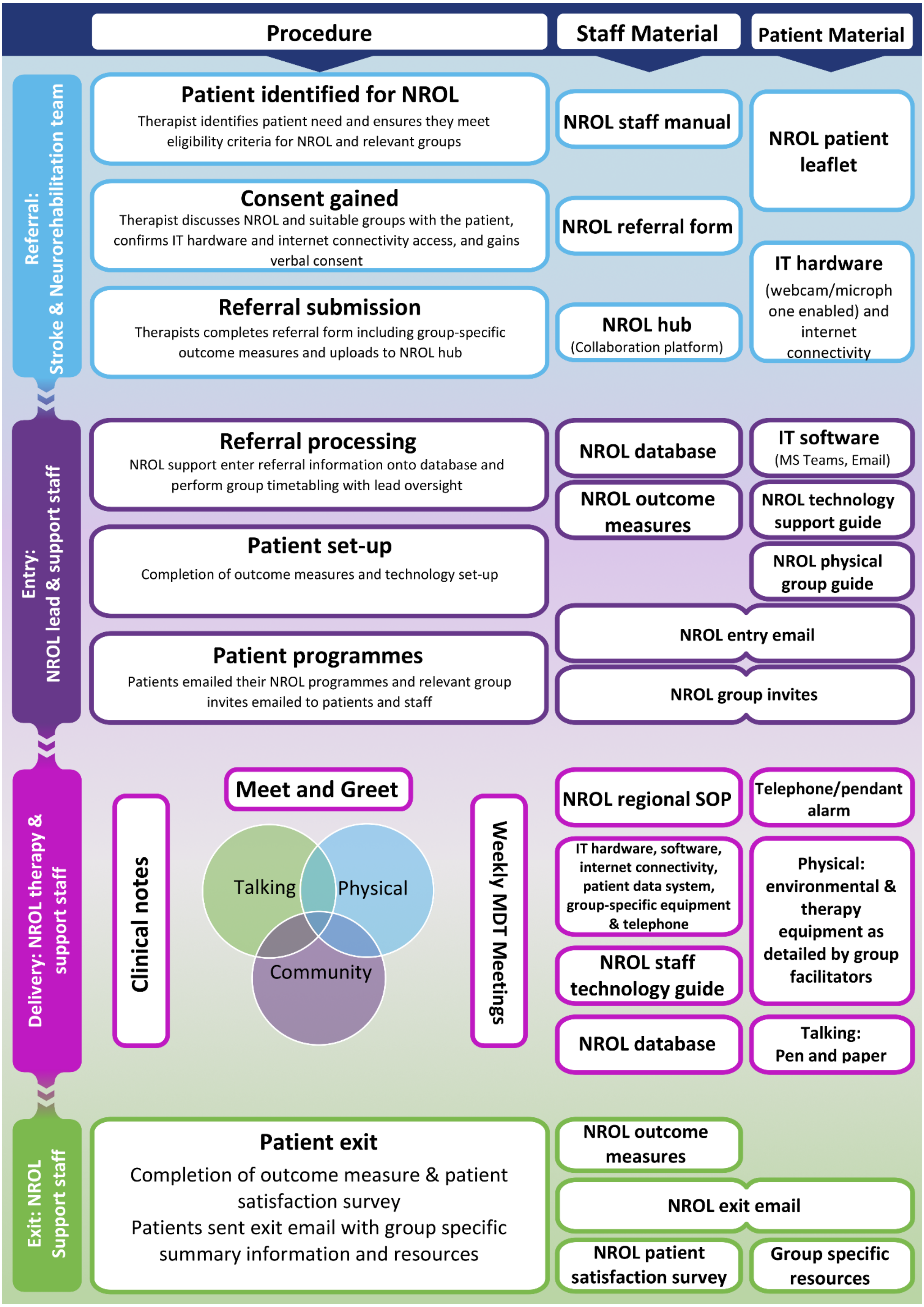

### CONSENTING A PATIENT FOR NROL

After identifying a suitable patient, therapists should discuss NROL with the patient, including providing a description of their relevant group(s). The NROL patient leaflet can be provided. Eligibility should also be confirmed against the group-specific criteria described in this manual. If a patient is able and willing to participate in NROL, they must provide verbal consent which is then recorded on the NROL referral form.

**To take part in NROL a patient needs to consent to:**

- participating in group-based online therapy from NHS partner trusts with other NHS staff able to observe
- agree to keep their camera on throughout sessions for safety (sessions are not recorded)
- consent to sharing their email address with NROL staff for NROL communication purposes

Etiquette and expectations regarding attendance, respect, and privacy should be outlined when discussing referral to NROL, are outlined in written documentation emailed to the patient before their NROL programme begins and at the initial 'NROL Meet and Greet session'.

### REFERRING A PATIENT TO NROL

Patients receiving active care from a stroke or neuro rehab team can be referred to NROL by a treating therapist. NROL referral forms are available from the NROL Hub MS Teams site (for access). The form collects patients' essential personal, medical, and current rehabilitation details, next of kin contact details, IT device details, and provides space for indicating the groups referred to. The form also provides space to document consents and response for any group-specific outcome measures. Once completed, the form is re-uploaded to the NROL Hub and then processed by the NROL team. A step-by-step guide is provided.

**To make a referral:**

*1. Get the latest referral form*

- Go to the '**NROL Hub MS** teams' general site and click on the '**Files**' tab at the top of the page
- Right click on the '***TEMPLATE_NROL referral form***' file pinned at the top and right click and select '**Download**'

*2. Download, save locally and complete*

- Open the downloaded referral form and **save** on your local drive using the following naming structure: **Patient initials_NROL Referral_Block number**
- Make sure to fill out **ALL** fields, then re-save

3. *Re-upload completed form to NROL Hub*

- On the NROL Hub (in elht.nhs.uk Teams), select your trusts folder, then select the '**Files'** tab and the '**New referrals**' folder
- Select '**Upload'** at the top of the page and select '**Files'** from the dropdown menu. Select the referral form you just completed that is saved on your local drive and then click '**Open**' at the bottom.
- The new referral form document with patient initials will now be uploaded, once it is processed it will be moved by NROL admin to the NROL Delivery channel which has limited access.

### GROUPS OVERVIEW

Groups running in each NROL block can differ and are determined by consensus based on patient need. Suggestions can be made for new groups to meet patients' needs.

*Groups run to date include:*

**Community**

Community groups are offered to all patients and include an introduction to NROL (Meet and Greet) and an optional weekly peer support group (Café NROL).

**Targeted therapy**

Targeted therapy groups can include 'physical' groups involving exercise or upper limb rehabilitation led by physiotherapy and occupational therapy staff, and 'talking' groups including cognitive rehabilitation, living well, fatigue management or communication therapy, led by occupational therapy, psychology and speech and language therapy staff. Many of the groups are jointly led by more than one discipline. Targeted therapy groups encompass interactive, educational, and practical elements.

| | | |
|---|---|---|
| *Community* | Meet and Greet | NROL programmes start with a session to introduce NROL and the therapy team, and provide an opportunity to test out the Teams technology |
| | Café NROL | An inclusive community group to provide an opportunity for peer support and group discussion |
| *Physical* | NROL with it | Involves exercise to improve strength, flexibility, mobility, balance, and fitness |
| | Upper limb group | Involves activities to increase function of the upper limb |
| *Talking* | Cogs in Motion | Aims to support people with their cognitive functioning and includes education and activities covering topics such as brain anatomy, attention, information processing and memory |
| | Cogs in Action | A practical group for patients with cognitive and/or cognitive communication difficulties. Run jointly by OT and SLT. All patients receive peer support from each other which helps their motivation and acceptance of their difficulties. Patients with overall cognitive difficulties need to have completed the Cogs in Motion group first. |
| | Simply Speaking | For patients with dysarthria, focusing on strategies to improve speech intelligibility |
| | Tip of the Tongue | Supports patients who have aphasia, and specifically word finding difficulties |
| | Living Well | Provides an opportunity to think about how we can live a good life, even with health issues, how we can keep focused and moving forward |
| | Fatigue Management | Aims to give information regarding fatigue, looking at how and why it affects us and provide practical strategies that can be implemented |

### MEET AND GREET: Community group

| Description | NROL starts with an 'NROL Meet and Greet' community session. The aim of this one-off session is to manage expectations, deal with any queries or concerns, provide an opportunity to practice using the Microsoft Teams platform and outline online group etiquette e.g., confidentially, respectful and active engagement in the sessions. Each group is described and the NROL team are introduced. |
|---|---|

**Patient Eligibility**: All patients joining the NROL programme for the first time. Patients must be suitable for NROL.

**Patient Equipment and Setting:**

- Patients participate from home using their own tablet/PC/laptop with webcam and microphone, and internet access.
- NROL Technology assistance is provided for initial NROL set-up and within sessions
- Patients should have a telephone in case of technology issues or adverse events and should wear their pendant alarm if they have one
- Patients should ensure they are in a quiet, private room when joining the group

**Staffing and Facilitation:**

- Sessions are delivered by at least two NROL therapy staff members, supported by a patient volunteer and with student involvement as appropriate
- NROL group facilitators
  - deliver sessions remotely from their work base, which might be different for each of them
  - require a webcam enabled PC/laptop/tablet, internet access, and MS Teams installed, headsets recommended
  - require access to the NROL database for accessing patient details, timetabling information and recording attendance, and a telephone in case of adverse events
  - must ensure they are in a private, well-lit, quiet space and ideally wearing an NROL t-shirt if delivering
  - coordinate a clinical notes entry for all patients after each session
  - can access NROL technology support as needed

**Session Format**

- Patients are welcomed and the purpose and structure of NROL is discussed, with PowerPoint presentations, described below, to complement. Facilitated discussion addresses general queries or concerns.

**Session Materials**

- PowerPoint presentations supplement facilitated conversations including slides on 'What is NROL', 'Meet the Team' and 'Group etiquette'.

### CAFÉ NROL: Community group

| Description | |
|---|---|
| Description | An optional community group for everyone participating in NROL. This group provides an opportunity for peer support and group discussion. It aims to encourage self-management and provide an opportunity to consolidate targeted therapy session, an open floor for patients to express themselves, a platform to share experiences and strategies, and a chance to learn about brain injury and neurological conditions. 'Café NROL' usually involves a 1-hour session for each week within the 6-week NROL block. |

**Patient Eligibility:** All patients joining the NROL programme are invited to the 'Café NROL' group. Patients must be suitable for NROL.

**Patient Equipment and Setting:**

- Patients participate from home using their own tablet/PC/laptop with webcam and microphone, and internet access
- NROL technology assistance is provided for initial NROL set-up and within sessions
- Patients should have a telephone in case of technology issues or adverse events and should wear their pendant alarm if they have one
- Patients should ensure they are in a quiet, private room when joining the group

**Staffing and Facilitation:**

- Sessions are delivered by at least two NROL therapy staff members, supported by a patient volunteer and with student involvement as appropriate
- NROL group facilitators
  - deliver sessions remotely from their work base, which might be different for each of them
  - require a webcam enabled PC/laptop/tablet, internet access, and MS Teams installed, headsets recommended
  - require access to the NROL database for accessing patient details, timetabling information and recording attendance, and a telephone in case of adverse events
  - must ensure they are in a private, well-lit, quiet space and ideally wearing an NROL t-shirt if delivering
  - coordinate a clinical notes entry for all patients after each session
  - can access NROL technology support as needed

**Session Format:**

- Facilitated conversation and group discussion incorporating professional advice and patient's perspectives. PowerPoint presentations, described below, to complement.

**Session Materials:**

- PowerPoint presentations to supplement facilitated conversation including slides on neuroplasticity and brain function, fatigue/pacing, memory, acceptance, physical activities and health choices, useful organisations/contact

### NROL WITH IT: Physical targeted therapy group

| | |
|---|---|
| **Description** | This physical therapy group involving exercise to improve strength, flexibility, mobility, balance and fitness. Patients are observed and advised on technique. 'NROL with it' usually involves a 45-minute session for each week within the 6-week NROL block. Maximum group size of about 12 patients. |

**Patient Eligibility**: Patients are identified by their treating therapists and referred via the NROL referral form. Patients must be suitable for NROL. Patients must also meet the following group specific criteria.

| | |
|---|---|
| **Inclusion** | Under the active care of a PT<br>Able to complete 30 mins of exercise (seated/ standing or a combination of both)<br>Can follow basic instructions with or without carer support (if requires carer they will need to be available during the remote session)<br>Can sustain attention for at least 30 minutes at a time<br>Can stand with support (family or carer) or walking aid which can be used during the sessions<br>Can communicate adequately in English, with support if needed<br>Can commit to attending and be willing to engage in 6 sessions |
| **Exclusion** | Unstable cardiac issues, recent DVT, or severe breathing difficulties<br>Any conditions which require active monitoring during exercise e.g., for consistent low or high BP, shortness of breath at rest, medical instability |

**Patient Equipment and Setting**:

- Patients participate from home using their own tablet/PC/laptop with webcam and microphone, and internet access
- NROL Technology assistance is provided for initial NROL set-up and within sessions
- Patients should have a telephone in case of technology issues or adverse events and should wear their pendant alarm if they have one
- Patients should ensure they are in a quiet, private room when joining the group
- Patients are advised to sit on a stable chair, preferably with arms
- Patients should have their usual walking aid or a sturdy chair/table in front or to the side of their chair
- Patients advised to wear loose comfortable clothing for exercise and flat shoes, preferably trainers
- Patients should have a drink within reach

**Staffing and Facilitation**:

- Sessions are delivered by at least two neurological PTs, with support from assistant practitioner or a student if available
- NROL group facilitators
  - deliver sessions remotely from their work base, which might be different for each of them
  - require a webcam enabled PC/laptop/tablet, internet access, and MS Teams installed, headsets recommended
  - require access to the NROL database for accessing patient details, timetabling information and recording attendance, and a telephone in case of adverse events
  - must ensure they are in a private, well-lit, quiet space and ideally wearing an NROL t-shirt if delivering
  - coordinate all clinical notes entry for all patients after each session
  - can access NROL technology support as needed

**Session Format:**

- Session content is flexible and adapted/tailored to patient need/ability (e.g., different balance positions) and can include functional strengthening, endurance exercise, balance training, sitting or standing, guided use of support/aid, sit to stand challenge

**Session Materials:**

- Patients are provided with information about camera set up and advice on clothing and footwear
- Staff crib sheet with breakdown of exercises for each session used by therapists. Set up with programme of exercises as a reference

### UPPER LIMB: Physical targeted therapy group

| | |
|---|---|
| **Description** | The aim of this group is to promote upper limb movement, function and care. Patients are observed and advised on technique. The 'Upper Limb' group usually involves a 1-hour session for each week within the 6-week NROL block. Maximum group size of about 10 patients. |

**Patient Eligibility**: Patients are identified by their treating therapists and referred via the NROL referral form. Patients must be suitable for NROL. Patients must also meet the following group specific criteria.

| | |
|---|---|
| **Inclusion** | Under the active care of a PT or OT<br>Have an upper limb impairment due to acquired brain injury, stroke, or neurological diagnosis<br>Can follow basic instructions with or without carer support (if requires carer they will need to be available during the remote session)<br>Can sustain attention for at least 30 minutes at a time<br>Can communicate adequately in English, with support if needed<br>Can commit to attending and be willing to engage in 6 sessions |
| **Exclusion** | Freezing or frozen stage adhesive capsulitis<br>Acute shoulder/upper limb injury (including but not exclusive to fractures, dislocations, recent surgery, lacerations/open or newly healing wounds)<br>Unstable CVS or recent DVT |

**Patient Equipment and Setting**:

- Patients participate from home using their own tablet/PC/laptop with webcam and microphone, and internet access
- NROL Technology assistance is provided for initial NROL set-up and within sessions
- Patients should have a telephone in case of technology issues or adverse events and should wear their pendant alarm if they have one
- Patients should ensure they are in a quiet, private room when joining the group
- Patients are advised to sit on a stable chair at a table
- Patients advised to wear loose comfortable clothing
- Equipment determined prior to each session, patient may need for example small weight, moisturising cream for sensory re-education, functional items e.g. cup, towel, pegs

**Staffing and Facilitation**:

- Sessions are delivered by at least two PT/OTs, with support from assistant practitioner or a student if available
- NROL group facilitators
  - deliver sessions remotely from their work base, which might be different for each of them
  - require a webcam enabled PC/laptop/tablet, internet access, and MS Teams installed, headsets recommended
  - require access to the NROL database for accessing patient details, timetabling information and recording attendance, and a telephone in case of adverse events
  - must ensure they are in a private, well-lit, quiet space and ideally wearing an NROL t-shirt if delivering
  - coordinate a clinical notes entry for all patients after each session
  - can access NROL technology support as needed

**Session Format:**

- Warm up, followed by stretches, strength exercises, sensory re-education and functional tasks
- Mental imagery is incorporated
- Exercises and activities are graded and adapted depending on the patient needs

**Session Materials:**

- The group facilitators have a crib sheet which follows the basic programme

### COGS IN MOTION: Talking targeted therapy group

| | |
|---|---|
| Description | This group aims to help patients understand their cognitive function and ways to manage cognitive difficulties. It includes education and activities covering topics such as brain anatomy, attention, information processing and memory. 'Cogs in Motion' usually involves a 1-hour session for each week within the 6-week NROL block. Maximum group size of about 6 patients. |

**Patient Eligibility**: Patients are identified by their treating therapists and referred via the NROL referral form. Patients must be suitable for NROL. Patients must also meet the following group specific criteria.

| | |
|---|---|
| Inclusion | Under the active care of an OT<br>Have a cognitive difficulty resulting from an acquired brain injury, stroke, or neurological diagnosis<br>Can sustain attention for at least 5 minutes at a time and be able to follow basic instructions<br>Can communicate adequately in English, with support if needed<br>Can commit to attending and be willing to engage in all 6 sessions |
| Exclusion | Patients with moderate to severe expressive and/or receptive language difficulties, please discuss with a SLT if required<br><br>Patients with palliative conditions or neurodegenerative conditions |

**Patient Equipment and Setting**:

- Patients participate from home using their own tablet/PC/laptop with webcam and microphone, and internet access
- NROL technology assistance is provided for initial NROL set-up and within sessions
- Patients should have a telephone in case of technology issues or adverse events and should wear their pendant alarm if they have one
- Patients should ensure they are in a quiet, private room when joining the group
- Patients are encouraged to have with them a notepad and pen to allow note taking

**Staffing and Facilitation**:

- Sessions are delivered by at least one OT with expertise in cognitive therapy, with support from either a second OT, an assistant practitioner, or a student.
- NROL group facilitators
  - deliver sessions remotely from their work base, which might be different for each of them
  - require a webcam enabled PC/laptop/tablet, internet access, and MS Teams installed, headsets recommended
  - require access to the NROL database for accessing patient details, timetabling information and recording attendance, and a telephone in case of adverse events
  - must ensure they are in a private, well-lit, quiet space and ideally wearing an NROL t-shirt if delivering
  - coordinate a clinical notes entry for all patients after each session
  - can access NROL technology support as needed

**Session Format**:

- Sessions involve structured questions to generate discussion on areas of cognition, alongside multiple-choice quizzes, and interactive tasks to demonstrate the cognitive skill being discussed that week
- Sessions include; Week 1 – Introduction session with ice breakers, Week 2 – 'All about the brain', Week 3 – Attention skills, Week 4 – Information processing, Week 5 – Memory skills, Week 6 – Executive skills

**Session Materials:**

- Patients receive an educational information pack at week 1, which includes 5 booklets for patients to read prior to each group. They also receive a 'next steps' leaflet on completion of the program.
- Copies of the booklets, session crib sheets and the session activities are accessed from the NROL Hub.

### COGS IN ACTION: Talking targeted therapy group

| | |
|---|---|
| Description | This is a practical group for patients with cognitive and/or cognitive communication difficulties, with a focus on practicing strategies to improve on their difficulties. 'Cogs in Action' usually involves five 1-hour sessions within the 6-week NROL block. Maximum group size of about 6 patients. |

**Patient Eligibility**: Patients are identified by their treating therapists and referred via the NROL referral form. Patients must be suitable for NROL. Patients must also meet the following group specific criteria.

| | |
|---|---|
| Inclusion | Under the active care of an OT or SLT<br>Have a cognitive difficulty and/or cognitive communication difficulty following an acquired brain injury, stroke, or neurological diagnosis<br>Patients with cognitive difficulties need to have completed the NROL 'Cogs in Motion' sessions first and read the information packs accompanying those sessions<br>Patients with cognitive communication difficulties do not necessarily need to have attended the Cogs in Motion sessions BUT <u>any</u> referral to this group needs to have been discussed with the SLTs leading the group first<br>Can sustain attention for at least 5 minutes at a time and be able to follow basic instructions<br>Can communicate adequately in English, with support if needed<br>Can commit to attending and be willing to engage in all 5 sessions |
| Exclusion | Patients with moderate to severe expressive and/or receptive language difficulties, please discuss with a SLT if required<br>Patients with palliative conditions or neurodegenerative conditions |

**Patient Equipment and Setting**:

- Patients participate from home using their own tablet/PC/laptop with webcam and microphone, and internet access
- NROL Technology assistance is provided for initial NROL set-up and within sessions
- Patients should have a telephone in case of technology issues or adverse events and should wear their pendant alarm if they have one
- Patients should ensure they are in a quiet, private room when joining the group
- Patients are encouraged to have with them a notepad and pen to allow note taking and for use as a memory strategy

**Staffing and Facilitation**:

- Sessions are delivered by an OT or SLT with expertise in cognitive therapy with support from either a second OT/SLT, an assistant practitioner, or a student.
- NROL group facilitators
    - deliver sessions remotely from their work base, which might be different for each of them
    - require a webcam enabled PC/laptop/tablet, internet access, and MS Teams installed, headsets recommended
    - require access to the NROL database for accessing patient details, timetabling information and recording attendance, and a telephone in case of adverse events
    - must ensure they are in a private, well-lit, quiet space and ideally wearing an NROL t-shirt if delivering
    - coordinate a clinical notes entry for all patients after each session
    - can access NROL technology support as needed

**Session Format:**

- Each session covers different strategies to manage cognitive difficulties (e.g. problems with attention, memory, information processing etc). If patients with cognitive communication difficulties are attending, discussion is held over how these cognitive difficulties impact on communication and again strategies are practised to manage this.
- Sessions start with a general check-in and homework review, followed by an explanation of the strategy under focus and an interactive session with opportunities to practise using this strategy. The last 10 minutes is used to recap the session, answer patient questions and set any homework.

### SIMPLY SPEAKING: Talking targeted therapy group

| Description | |
|---|---|
| | This group is for patients with mild-moderate dysarthria who could benefit from strategies to improve speech intelligibility. 'Simply Speaking' usually involves a 1-hour session for each week within the 6-week NROL block. Maximum group size of about 6 patients. |

**Patient Eligibility**: Patients are identified by their treating therapists and referred via the NROL referral form. Patients must be suitable for NROL. Patients must also meet the following group specific criteria.

| Inclusion | |
|---|---|
| | Under the active care of a SLT or had it recently and needs a 'refresher'<br>Comprehension and cognition must be at a level where they can participate without an SLT being physically present and follow verbal instructions<br>Can sustain attention for at least 15 minutes at a time and participate for at least 30 minutes of the session<br>Can communicate adequately in English, with support if needed<br>Can commit to attending and be willing to engage in 6 sessions |

**Patient Equipment and Setting**:
- Patients participate from home using their own tablet/PC/laptop with webcam and microphone, and internet access
- NROL Technology assistance is provided for initial NROL set-up and within sessions
- Patients should have a telephone in case of technology issues or adverse events and should wear their pendant alarm if they have one
- Patients should ensure they are in a quiet, private room when joining the group
- Patients are encouraged to have with them a notepad and pen to allow note taking

**Staffing and Facilitation**:
- Sessions are delivered by at least one SLT, with support from either a second SLT, an assistant practitioner, or a student(s) supervised by the SLT
- NROL group facilitators
    - deliver sessions remotely from their work base, which might be different for each of them
    - require a webcam enabled PC/laptop/tablet, internet access, and MS Teams installed, headsets recommended
    - require access to the NROL database for accessing patient details, timetabling information and recording attendance, and a telephone in case of adverse events
    - must ensure they are in a private, well-lit, quiet space and ideally wearing an NROL t-shirt if delivering
    - coordinate a clinical notes entry for all patients after each session
    - can access NROL technology support as needed

**Session Format**

- Sessions start with a general check-in and homework review, followed by an explanation of the strategy under focus and an interactive session (determined by their impairment level) to practise using this strategy. The last 10 minutes is used to recap the session, answer patient questions and set any homework.
- Each session covers a different strategy: diaphragmatic breathing, over articulation, chunking (i.e, breaking words/sentences down into manageable chunks), pacing, all with a focus of using these strategies functionally.

**Session Materials**
- Crib sheets, session activities are accessed from the NROL Hub
- Used during session – PowerPoint presentations

### TIP OF THE TONGUE: Talking targeted therapy group

| | |
|---|---|
| **Description** | This group is designed to support patients with aphasia and specifically with word finding difficulties. 'Tip of the Tongue' usually involves five 1-hour sessions within the 6-week NROL block. Maximum group size of about 6 patients. |

**Patient Eligibility**: Patients are identified by their treating therapists and referred via the NROL referral form. Patients must be suitable for NROL. Patients must also meet the following group specific criteria.

| | |
|---|---|
| **Inclusion** | Must be under the active care of a SLT or had it recently and needs a 'refresher'<br>Comprehension and cognition must be at a level where they can participate without an SLT being physically present and follow verbal instructions<br>Can sustain attention for at least 15 minutes at a time and participate for at least 30 minutes of the session<br>Can communicate adequately in English, with support if needed<br>Can commit to attending and be willing to engage in 5 sessions |

**Patient Equipment and Setting**:

- Patients participate from home using their own tablet/PC/laptop with webcam and microphone, and internet access
- NROL Technology assistance is provided for initial NROL set-up and within sessions
- Patients should have a telephone in case of technology issues or adverse events and should wear their pendant alarm if they have one
- Patients should ensure they are in a quiet, private room when joining the group
- Patients are encouraged to have with them a notepad and pen to allow note taking, if they are able to do so

**Staffing and Facilitation**:

- Sessions are delivered by at least one SLT, with support from either a second SLT, an assistant practitioner, or a student(s) supervised by the SLT
- NROL group facilitators
  - must have knowledge about the strategies and how they are to be used, knowledge about aphasia and word finding difficulties
  - deliver sessions remotely from their work base, which might be different for each of them
  - require a webcam enabled PC/laptop/tablet, internet access, and MS Teams installed, headsets recommended
  - require access to the NROL database for accessing patient details, timetabling information and recording attendance, and a telephone in case of adverse events
  - must ensure they are in a private, well-lit, quiet space and ideally wearing an NROL t-shirt if delivering
  - coordinate a clinical notes entry for all patients after each session
  - can access NROL technology support as needed

**Session Format:**

- Sessions start with a general check-in and homework review, followed by an explanation of the strategy under focus and an interactive session (determined by their impairment level) to practise using this strategy. The last 10 minutes is used to recap the session, answer patient questions and set any homework.
- Sessions 1-4 cover different strategies (non-verbal communication, circumlocution and semantic cues, synonyms and phonemic and graphemic cues). Session 5 is an opportunity to bring these strategies together via role play.

**Session Materials**

- Crib sheets, session activities, information/therapy task sheets are accessed from the NROL Hub. The latter materials are aphasia friendly and can be shared online during the group or can be sent out via email/post depending on the group's preference.
- Used during session – PowerPoint presentations, any task sheets set as homework

### LIVING WELL: Talking targeted therapy group

| | |
|---|---|
| **Description** | The ‘Living Well’ group is a place to think about how we can live a good life, even with health issues, and how we can keep focused and moving forward. The group is a space to share ideas and coping strategies and make plans. Living Well usually involves three 1-hour sessions within the 6-week NROL block. Maximum group size of about 10 patients. |

**Patient Eligibility**: Patients are identified by their treating therapists and referred via the NROL referral form. Patients must be suitable for NROL including being under the active care of a stroke or neurorehabilitation team. Patients must also meet the following group specific criteria.

| | |
|---|---|
| **Inclusion** | Must consent to have detailed discussions about their rehabilitation journey and goals with other participants present. NB sometimes difficult feelings and thoughts are discussed but no-one is forced to share.<br>Can sustain attention for at least 5 minutes at a time and be able to engage in a group conversation<br>Can communicate adequately in English, with support if needed<br>Can commit to attending and be willing to engage in 3 all sessions |
| **Exclusion** | Patients with moderate to severe cognitive or expressive and/or receptive language difficulties, please discuss with an OT and/or SLT if required<br>Current significant risk of self-harm<br>Acutely distressed patient likely requiring one-one input<br>Significant acute distress triggered by hearing about others’ challenging circumstances |

**Patient Equipment and Setting**:
- Patients participate from home using their own tablet/PC/laptop with webcam and microphone, and internet access
- NROL Technology assistance is provided for initial NROL set-up and within sessions
- Patients should have a telephone in case of technology issues or adverse events and should wear their pendant alarm if they have one
- Patients should ensure they are in a quiet, private room when joining the group
- Patients are encouraged to have with them a notepad and pen to allow note taking

**Staffing and Facilitation**:
- Sessions are facilitated by a clinical psychologist with expertise in working with people with stroke/other neurological conditions, with either an assistant practitioner or a student supporting delivery
- NROL group facilitators
  - deliver sessions remotely from their work base, which might be different for each of them
  - require a webcam enabled PC/laptop/tablet, internet access, and MS Teams installed, headsets recommended
  - require access to the NROL database for accessing patient details, timetabling information and recording attendance, and a telephone in case of adverse events
  - must ensure they are in a private, well-lit, quiet space and ideally wearing an NROL t-shirt if delivering
  - coordinate a clinical notes entry for all patients after each session
  - can access NROL technology support as needed

**Session Format:**
- Session content is flexible according to issues raised by the group, but is broadly delivered as three sessions: 1) the challenges of living well with long-term health issues using a values-based approach; 2) goal-setting using values-based goals; 3) identifying and managing difficult thoughts and feelings
- The sessions focus on group discussion primarily, rather than formal didactic teaching, with the facilitator drawing out and making explicit the key learning points that emerge in the discussion, and relating the discussion back to the key concepts and skills. Experiential exercises are also sometimes used.
- Previous sessions and homework are reviewed as needed, and new homework activities are set.

**Session Materials:**

- Resources are provided summing up the key points, and including a values identification exercise, goal-setting and planning sheets, mindfulness and cognitive de-fusion exercises, and a coping and contingency plan. The Choice Point model is also provided for information.

### FATIGUE MANAGEMENT: Talking targeted therapy group

| | |
|---|---|
| **Description** | This group is designed to educate individuals about fatigue following a neurological condition, and to examine how and why it affects people. There are discussions about the different types of fatigue, and the factors that contribute. During the sessions practical coping strategies are discussed and practiced. The 'Fatigue Management' group usually involves a 1-hour session for each week within the 6-week NROL block. Maximum group size of about 10 patients. |

**Patient Eligibility**: Patients are identified by their treating therapists and referred via the NROL referral form. Patients must be suitable for NROL including being under the active care of a stroke or neurorehabilitation team. Patients must also meet the following group specific criteria.

| | |
|---|---|
| **Inclusion** | A willingness to engage and discuss their own experiences during the session<br>Can sustain attention for at least 5 minutes at a time and follow basic instructions<br>Can communicate adequately in English, with support if needed<br>Can commit to attending and be willing to engage in 6 sessions |
| **Exclusion** | Patients with moderate to severe cognitive or expressive and/or receptive language difficulties, please discuss with an OT and/or SLT if required |

**Patient Equipment and Setting**:
- Patients participate from home using their own tablet/PC/laptop with webcam and microphone, and internet access
- NROL Technology assistance is provided for initial NROL set-up and within sessions
- Patients should have a telephone in case of technology issues or adverse events and should wear their pendant alarm if they have one
- Patients should ensure they are in a quiet, private room when joining the group
- Patients are encouraged to have with them a notepad and pen to allow note taking

**Staffing and Facilitation**:
- Sessions are facilitated by an OT with expertise in working with people with stroke/other neurological conditions, with either an assistant practitioner or a student supporting delivery
- NROL group facilitators
  - deliver sessions remotely from their work base, which might be different for each of them
  - require a webcam enabled PC/laptop/tablet, internet access, and MS Teams installed, headsets recommended
  - require access to the NROL database for accessing patient details, timetabling information and recording attendance, and a telephone in case of adverse events
  - must ensure they are in a private, well-lit, quiet space and ideally wearing an NROL t-shirt if delivering
  - coordinate a clinical notes entry for all patients after each session
  - can access NROL technology support as needed

**Session Format:**
- Sessions are education based with both didactic and interactive elements

**Session Materials:**
- Handouts are sent before the group – Values checklist and action plan, what empties my batteries, fatigue management assessment and fatigue diary (preference for paper copies).
- Handouts sent after the group – copy of PowerPoint, visualisation script, mindfulness script, pacing and boom & bust cycle, relaxation apps.
- Materials used during sessions – PowerPoint, relaxation scripts (visualisation, ACT book for mindfulness)
